# WHEN SEX MATTERS: A COMPARATIVE GENERALISED STRUCTURAL EQUATION MODEL (GSEM) FOR THE DETERMINANTS OF STUNTING AMONGST UNDER-FIVES IN UGANDA

**DOI:** 10.1101/2022.04.25.22274287

**Authors:** M Vallence Ngabo, Leonard Atuhaire, Peter Clever Rutayisire

## Abstract

The main aim of this study was to establish the differences in both the determinants of stunting and the causal mechanism through which the identified determinants influence stunting amongst male and female under-fives in Uganda. Literature shows that male children below the age of five years are at a higher risk of being stunted than their female counterparts. Specifically, studies in Uganda indicate that being a male child is positively associated with stunting while being a female is negatively associated with stunting.

Data for 904 male and 829 female under-fives was extracted form UDHS-2016 survey dataset. Key variables for this study were identified and used in generating relevant models and paths. Structural equation modeling techniques was used in its generalized form (GSEM). The generalized nature necessitated specifying both the family and link functions for each response variables in the system of the model. Sex of the child (b4) was used as a grouping factor and the height for age (HAZ) scores were used to construct the status for stunting of under-fives.

The estimated models and path clearly indicated that the set of underlying factors that influence male and female under-fives respectively were different and the path through which they influence stunting were different. However, some of the determinants that influenced stunting amongst male under-fives also influenced stunting amongst the female under-fives.

To reduce the stunting problem to the desirable state, it is important to consider the multifaceted and complex nature of the risk factors that influence stunting amongst the under-fives but more importantly consider the different sex specific factors and their causal mechanism or paths through which they influence stunting.

## Introduction

Low height-for-age in children also known as child stunting is one of the most important indicators of child’s wellbeing and health. The broad belief is that stunting just like other forms of child malnutrition is associated with household’s poor socioeconomic status, poor maternal health and nutrition during and after pregnancy, frequent illness of the child, inappropriate infants and young child feeding and care practices, unpleasant environmental conditions and child inherent factors. At a point of inception, a child’s life starts, as thus poor nutrition during pregnancy and early childhood may be a major cause for poor growth. Stunting may go a long way beyond the child’s height-for-age defects to cognitive impairments (1) and this situation puts them at a disadvantage of performing well during their education career and also leads to poor performance at work (2). The potentially irreparable physical and neurocognitive damage that accompanies stunted growth is a major obstacle to human development in later years of life (2). Stunting increases the frequency of illness and is likely to increase the risk of death (3). Moreover stunting is a cyclical process because women who were themselves stunted in childhood tend to have stunted offspring, creating an intergenerational cycle of poverty and reduced human capital that is difficult to break (4).

By the year 2020, the estimates for stunted under-fives stood at 149.2 million which accounted for 22% of all the children under the age of five years worldwide (5). It is a global public health concern and disparities in its distribution world over should be given attention. Continental, regional and country level variations do exist and this calls for different strategies and efforts to deal with the problem. Out of the 149.2 million stunted under-fives world over, 41% live in Africa while 53% live in Asia by the estimates of 2020 (5). Africa is a special case because as the number of child stunting is reducing elsewhere, stunting in Africa has actually increased from 54.4 million in 2000 to 61.4 million by 2020 (5). With estimates above 30%, East Africa is among the region with the highest levels of stunting in Africa (5) and this is not different with Uganda whereby about 3 in every 10 children are stunted.

Globally, sex of the child has been found to be one of the factors influencing stunting among the under-fives but the male children have had a higher risk of being stunted than the female children. Studies done in Sub-Saharan African mostly report a higher prevalence amongst male children than female children (6). In the food surplus region of Ethiopia, the odds of being stunted were more times for males than for females (7). Studies such as (8–11) done in Uganda mostly indicate that the sex of the child is among other factors with significancy in influencing stunting of the under-fives. Among the 6-59 months old’s in South-Western Uganda, boys were found to be 2.2 times more likely to be stunted than girls (9). Among the 6-59 months Karimojong children, it was found that male children were 1.8 times more likely to experience concurrent stunting and wasting compared with their females counterparts (12). There is little doubt that stunting is higher amongst the poor households than among the wealthier household but a closer look into the matter still reveals that boys were at a higher risk of being stunted than girls. More so, stunting was higher within the poorer households than their wealthier counterparts (13). Many other studies (6,14–16) support the fact that male children were more at the risk of being stunted than the female children. Just a few studies found a non-significant influence of sex of the child to stunting though the fact that boys were at a risk of being malnourished than girls was still evident (17). Based on the above literature, it appears that the risk of stunting amongst the male children while their female counterparts are not is not by chance and thus an investigation of the matter is worthwhile.

## Materials and methods

### The conceptual framework

The conceptual framework for this study is founded on the modified United Nations Children’s Education Fund (UNICEF) framework on the determinants of child malnutrition (18) which shows the structural system of possible path through which determinants operate to influence malnutrition in general and stunting in particular. Moreover, information from previous studies (19–23) identified the most influential factors of stunting amongst the under-fives was also used to shape better the current conceptual framework. Stunting, an indicator of undernutrition, is one of the four forms of malnutrition and one would not go wrong by using the malnutrition framework in the analysis of risk factors of stunting. The framework is clear on the fact that factors that influence the stunting are better categorized in different levels as; distal, intermediate, immediate and the child inherent factors.

Except for the child inherent factors that don’t have background determinants, all other category of factors were interrelated and happen to feed into each other at various levels. The distal factors may influence stunting of the child either directly or indirectly through either the intermediate or immediate factors. Just like the distal factors, the intermediate factors influence stunting either directly or indirectly through the immediate factors. The current study will also analyse the possibility of the interrelationship of factors within the same group because it possible for example household wealth index may influence mother’s education level and mother’s education level may be a factor in determining the current employment status.

According to the conceptual framework, the child inherent factor are the age and sex of the child. The distal factors include, those factors that determine the socioeconomic status of the household and they include; the education levels of the child’s parents or caregivers, mother’s employment status, household wealth index (a proxy of poverty or richness), region, place of residence and the religion. The intermediate factors which are the determinants of the environment and maternal health act as a medium to influence stunting and they include; mother’s age, mother’s weight (BMI), mother’s decision making (autonomy), parity in the household, antenatal timing and frequency, place of delivery, household membership, toilet facilities, sources of clean drinking water and health insurance coverage. The immediate causes of child stunting are basically child level and they include; breastfeeding initiation, dietary diversity, size and perceived weight at birth, birth order of the child, fever, cough and diarrhoea episodes within the two weeks preceding the interview. Accordingly, the conceptual framework shows the hierarchical nature of the risk factors where factors at one level feed into factors at other level(s) up to the overall independent variable. The conceptual framework also shows the fact that while stunting is the only purely endogenous variable, there are several other factors in the system that are either exogenous at one point or endogenous at another point (Fig 1)

**Figure 1.**
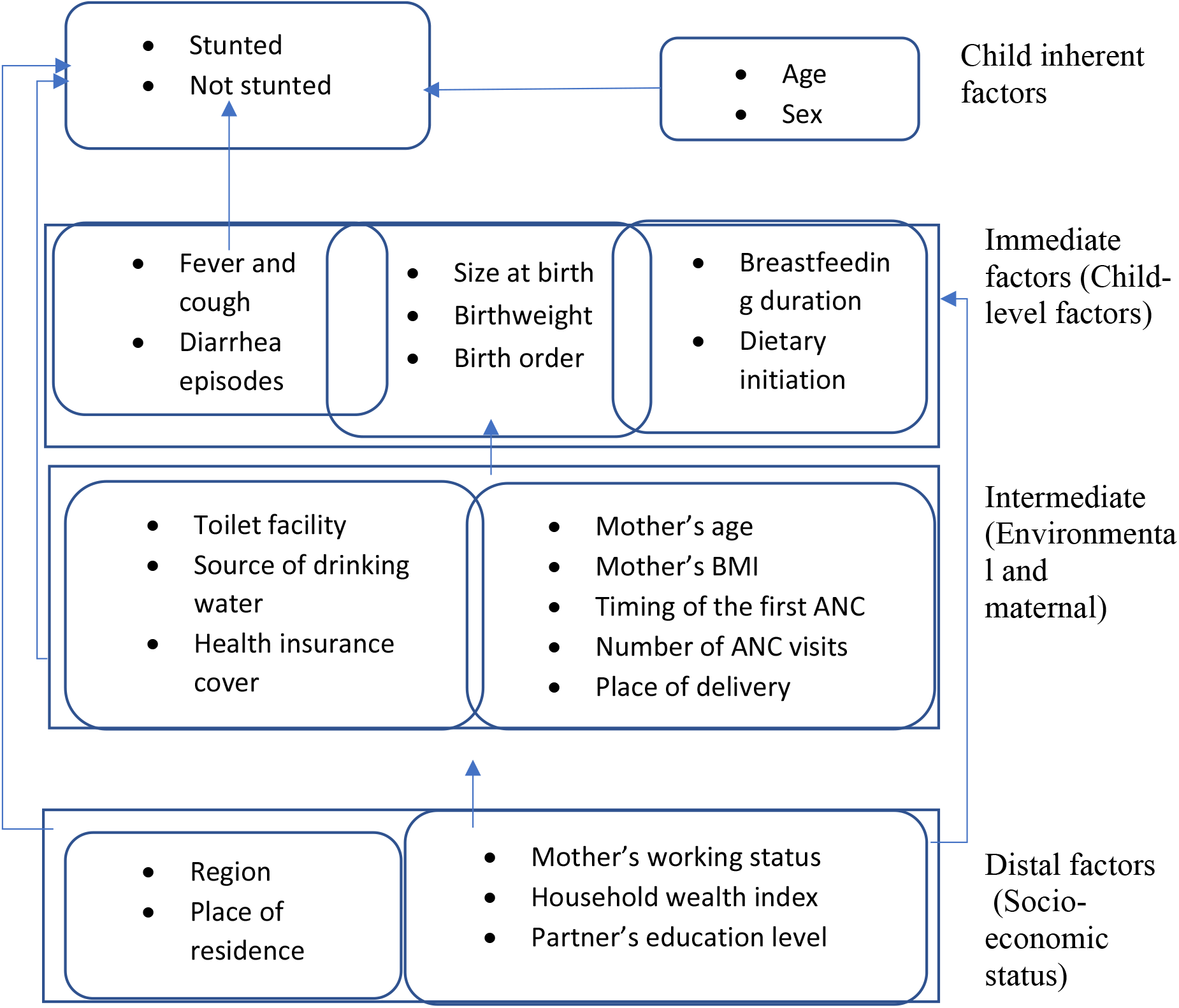
A conceptual framework for analyzing the risk factors of under-five Undernutrition (Adapted from Hein & Hoa, 2009; Mostafa, 2011; Nikoi & Anthamatten, 2014; Boah et al.,2019; United Nations Children’s Fund, 1991)

### Data and its sources

Data for this study was purely secondary data extracted from the child record (KR dataset) of the 2016 Uganda Demographic and Health Survey (2016 UDHS) (24). Approval for the use of the UDHS dataset was granted by ICF international via the online portal https://www.dhsprogram.com/. Cases for children aged 0-59 months whose anthropometric measures were successfully captured at the time of the survey were extracted from UDHS dataset and used in this study. UDHS survey was designed and implemented by Uganda Bureau of Statistics (UBOS) to provide estimates of the population and health indicators including fertility and child health for the country as a whole, rural and urban and for each of the 15 regions in Uganda (South Central, North Central, Busoga, Kampala, Lango, Acholi, Tooro, Bunyoro, Bukedi, Bugisu, Karamoja, Teso, Kigezi, Ankole, and West Nile). The survey employed a multistage and multisampling techniques whereby 15 clusters (regions) were selected at the first stage, households were selected at the second stage and finally interviews or appropriate measurements were carried out on household members who met the selection criteria. Overall, the survey used three types of questionnaires were used to capture information concerning household characteristics, fertility, morbidity, and child health were considered. All women in the selected households aged 15-49 were interviewed and anthropometric measurements as well as other relevant information for the children aged 0-59 month within the selected households were collected.

### Variable description

The purely endogenous variable for this study was stunting, a proxy for height-for-age z-scores that were less than -2standard deviations from the median of the reference population of children in the agegroup of 0-59 months old (25). These z-scores were calculated based on the 2006 child growth standards of the World Health Organization (WHO) (25). Observations whose z-scores values were outside the normal range of z-scores were flagged off. Accordingly, z-scores (*Zi*) were calculated as 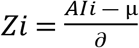 where *Ali* refer to the anthropometric indicator of i^th^ child, µ *and* ∂ respectively refer to the median and the standard deviation of the reference population. A z-score cutoff point (−2sd) was used to generate a dummy variable with binary outcomes for stunting

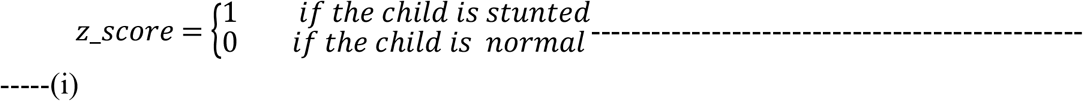

Independent variables were categorized as; child inherent, child level, immediate, intermediate and distal factors. Except the child inherent factors (sex and age) of the child that were purely exogenous, other variables in the system were either endogenous or exogenous because it was clear that some variables at one level were not only determinants of stunting but would also be influence other variables at the higher level in the system (Fig 1). Sex of the child was used as comparative variable since the study was following a comparative approach where analysis was made with respect to the sex of the child. Age of the child was categorized “0-5”, “6-11”, “12-23”, “24-35”, “36-47”, “48-59” month of age. Size of the child at birth (m18) was categorized into small and large while child wight at birth was categorized into underweight (<2.5kgs) normal (2.5<=bw<4 kgs) and high birth weight (>4kgs) at birth (26). Fever, malaria and diarrhoea episodes, birth order, duration of breast feeding, and initiation of complementary feeding were analyzed for possibility in influencing stunting of under-fives.

Intermediate level factors were considered as the possible determinants of the environment in which the child and the mother live. They included; mother’s age which was categorised and used as mother’s age-group (15-24; 25-34; 35-49), mothers’ weight (categorised as thin (BMI<18.5kgs/m_2_, normal (18.5<BMI<24.5kgs/m_2_) overweight (BMI>25kg/m_2_), place of delivery, household size, toilet facility, source of safe water, parity, mother’s autonomy and health insurance coverage, the timing of the first antenatal care (ANC), number of ANC visits. Given the fact that antenatal care services are free of charge in Ugandan government hospitals, the timing of ANC, the number of ANC visits during pregnancy and the place of delivery are major determinants of expectant mother’s health seeking behavior. In this study, distal factors were considered as factors that determine the socioeconomic status as well as the cultural factors of the household. They are considered very important in forming the basis of the welfare status of the household. They include; the spatial factor (region), place of residence (rural or urban), mother’s and father’s education level, mother’s employment and working status, religion and the wealth index of the household. The household wealth index is a composite measure of a household’s cumulative standards of living. Its calculated using easy-to-collect data on a household’s ownership of selected assets, such as televisions and bicycles; materials used for housing construction; and types of water access and sanitation facilities of the household. UDHS categorised households into wealth quintiles (lowest, second, middle, fourth and highest) as indicators of poorest to richest.

### Data analysis

The complex and multi-faceted nature of the problem of stunting and its risk factor presents a challenge in estimation. The system according to the conceptual framework (Fig, 1) reveals that a model that accommodates all variables in their statistical nature (categorical, count, continuous or discrete) is paramount. It was observed that the overall dependent variable (stunting) is binary in nature while other variables in the system were either ordinal, count, discrete or continuous yet they were to be used in the same model.

A generalized structural equation model (GSEM) was estimated. Since the study was comparative in nature, estimation of the model parameters involved sex of the child (b4) as a grouping variable. GSEM was chosen over other modeling techniques because of its ability to accommodate structural equations in a system that allows variables to feed into each other despite their nature and also because of its ability to relax the assumption of normality of variables, it accommodates both generalized linear responses as well as linear responses, it allows the use of factor variable notation (specifically in Stata), it allows for multi-level models and it is able to use more observations in the presence of missing values and it allows for group analysis. The distribution as well the link function that depends on the nature of a variable were specified for each of the endogenous variable in the system of simultaneous equations.

GSEMs represents a generalization of SEMs by allowing the use of all the types of variables and considering their distributions (27) as well as its advantages over other modeling approaches such as; (i) relaxation the assumption of normality of variables, (ii) it accommodates both generalized linear responses as well as linear responses, (iii) it allows the use of factor variable notation (specifically in Stata), (iv) it allows for multi-level models (v) it is able to use more observations in the presence of missing values and it allows for group analysis, make it a more preferred modeling approach in the current study.

### GSEM equation modelling

GSEM modeling approach combines the power and flexibility of both structural equation models (SEM) and generalized linear model (GLM) in a unified modeling framework (28). A combination of the statistical distribution of the family (*F*) and the link function g (·) for the response variable(s) were clearly specified within the structural modeling framework. Generally, let ley *yi* be a vector of endogenous variable, let *xi* be a vector of exogenous variables, and let ley *β′and ui* be the coefficient and error terms respectively, then *yi yi* = *β′xi* + *ui*, generalized to the linear model of the form *g*{*E*(*yi*)} = *β′xi* with *yi* ∼*F* (Fig. 2).

**Figure 2:**
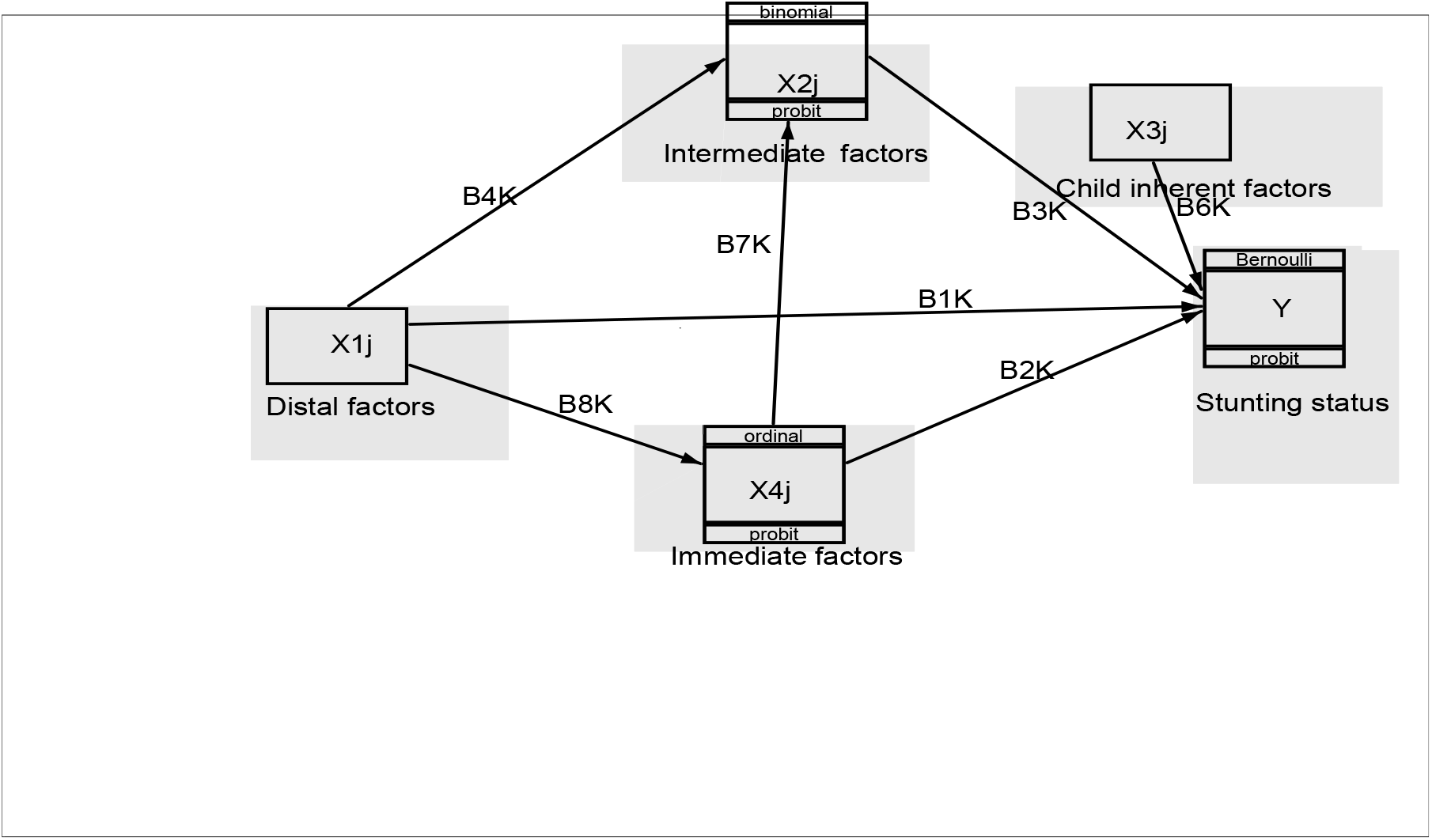
The generalized structural equation model; Note At each level the vector contains either count, discrete, continuous or ordinal data. The family and link descriptions may not carry much sense at this point int time.

Let *Yi* be the overall response variable (stunting status), *X*1*j, X*2*j, X*3*j, and X*4*j* be matrices for distal, immediate, intermediate and child inherent factors respectively. The direct effects of; distal, immediate, intermediate and child inherent on *Yi* as in Fig 2 were estimated as; *B*1*K, B*2*K, B*3*K, and B*7*K* respectively while the indirect effects were calculated as; *B*8*K* ∗ *B*2*K, B*4*K* ∗ *B*3*K, B*4*K* ∗ *B*6*K* ∗ *B*2*K, and B*8*K* ∗ *B*5*K* ∗ *B*3*K*. Let *BiK, i* = 1,2,3,…,8, be the effect of factors that feed into each other at different hierarchical levels, for example *B*4*K* is the effect of distal factor onto the immediate factors in the model (Fig 2). At each stage, the distribution and family of the response variable will be specified. An error term (ε) was generated of family Gaussian (quantitative and continuous) factors.

During the modelling process, it was considered important to avoid excessive number of parameters in the final output (appendix 1) and as such only important variables are reflected in the final model. Subsequently only parameters whose p-value <0.1 were maintained in the establishment and the analysis of the path model (Figs 3 and 4). Sex was specified as a grouping variable in generating comparative parameters for both male and female under-fives and analysis was possible with the use SEM/GSEM builder within STATA version 15.0

**Figure 3:**
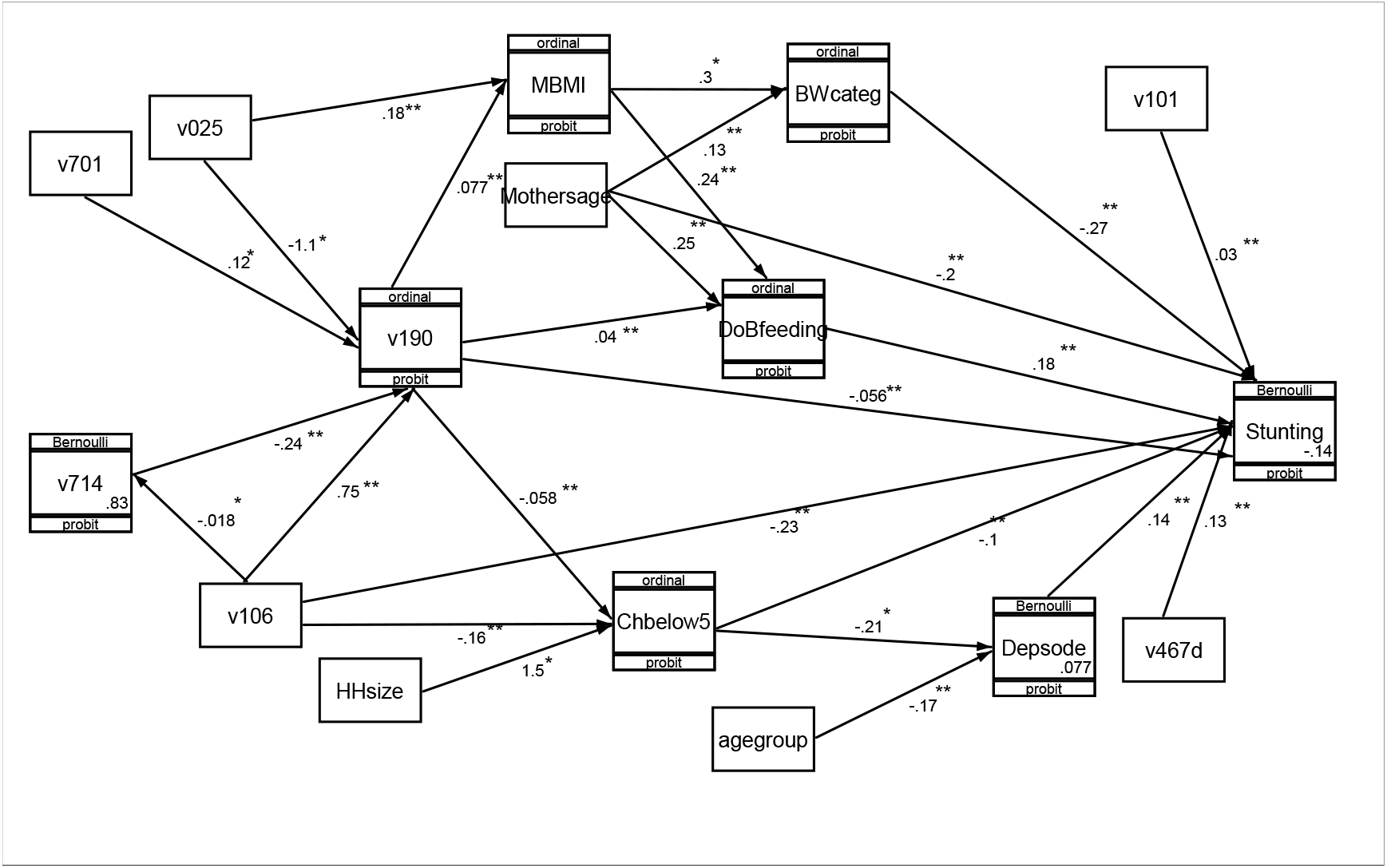
Path mechanism of statistically significant risk factors of male stunting

**Figure 4:**
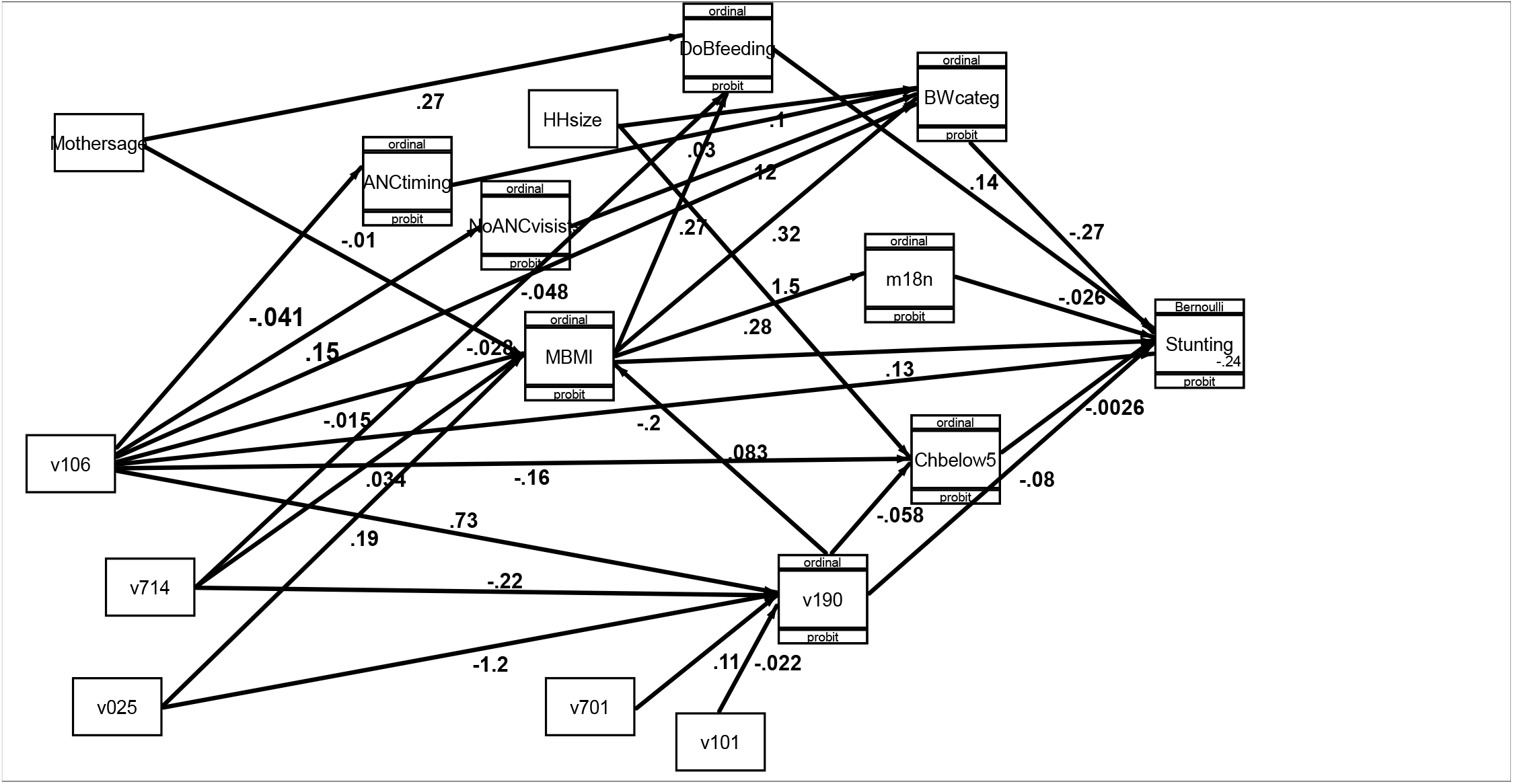
Path mechanism of statistically significant risk factors of female stunting

## Ethics statement

This paper is based on secondary data that exists in the public domain and is readily accessible upon request. Accordingly, the permission to access and use the dataset (UDHS 2016) was sought and granted via the DHS website (https://dhsprogram.com/data/dataset/Uganda_Standard-DHS_2016.cfm?flag=1). The DHS programme is a worldwide reputable organization for collecting and disseminating credible national representative data on health indicators especially data on household demographics and child nutrition. For the case of the Ugandan country team, the ICF institutional review board (IRB) reviewed and approved the UDHS 2016 tools for the survey as well as the procedure of obtaining written informed consent. The ICF IRB complied with the United States Department of Health and Human Services regulations for human subjects. Informed consent was obtained from participants in writing whereby a form was signed by the respondent as well, data on minors were collected on children based on mothers’ informed consent obtained in writing. Participants were given full information about the survey and their participation was purely voluntary. Confidentiality and keeping participants anonymous, elements were observed by exclusion of identifiers from the final dataset obtained. Further information regarding the conduct of the study that generated the data used can be found in the report of UDHS 2016 (24).

## Results

After realising that male and female under-five are affected differently by stunting, the major task in this study was to establish the likely reason why the status quo. The analysis was banked on GSEM modeling approach whereby, the path, the link and family of response variables in the GSEM structure were specified and a GSEM model was constructed and estimated. The estimation followed three levels; level one involved identification of significant risk factors, level two involved identification significant factors with respect to their levels while level three involved identification of the path which was pursued by identifying the determinants of significant determinants as identified in level 1 and 2. To avoid crowding of paths, the path diagram (Figs 3 and 4) only presented the path with respect to the significant risk factors of stunting. Parameters in the Tables 1 and 2 were generated for variables in combination without factor level specifications. The grouping variable was specified as sex of the child under-five years of age (b4) or 0-59 months and as such estimates were generated in comparisons of male child (Table 1) and female child (Table 2). Later analysis of the same model considered the specification of factor levels (Tables 3 and 4). Until this stage, the analysis targeted the identification of the most significant direct determinants of stunting with reference to the sex of the child. Path analysis was evoked by identifying the factors behind those determinants that were found to be significant at level one. Risk factors were considered statistically significant in case their p-value (p(z) was less or equal to 10%.

**Table 1:**
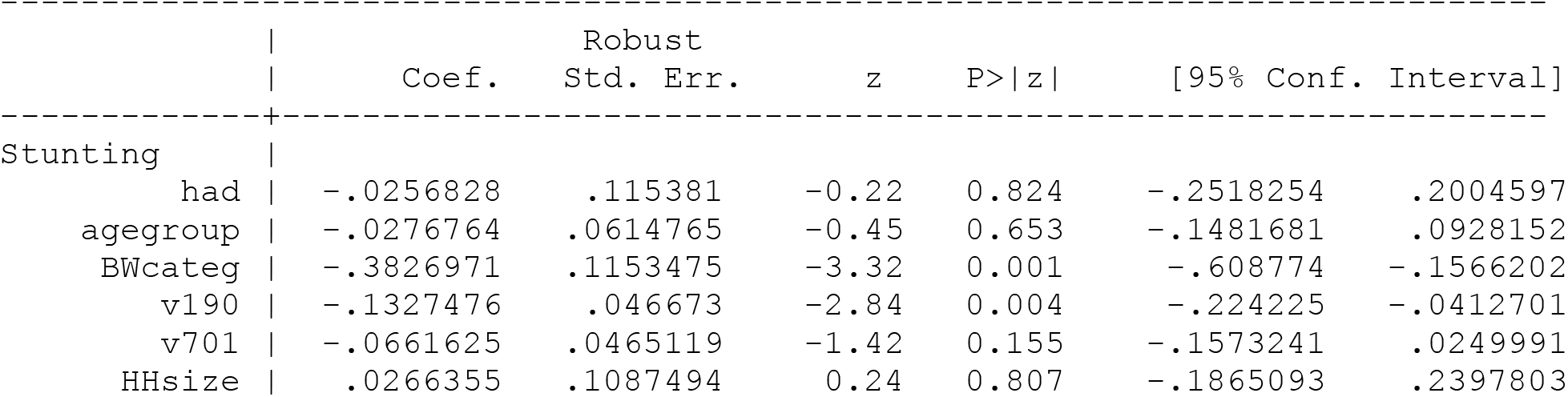
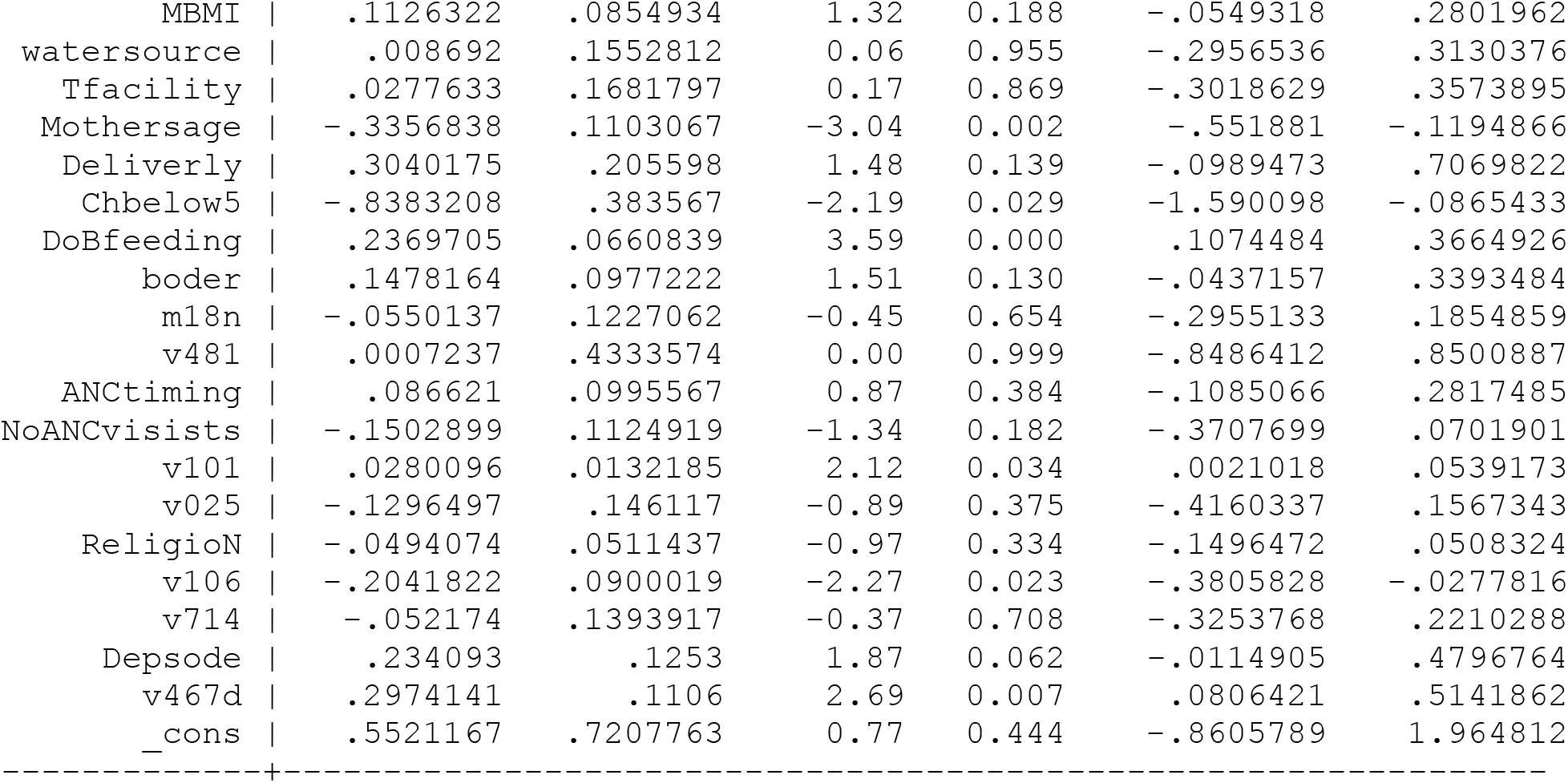
The risk factors responsible for stunting amongst the male under-fives (n=904)

|  |  | Coef. | Robust Std. Err. | z | P>\|z\| | [95% Conf. Interval] |  |
| --- | --- | --- | --- | --- | --- | --- | --- |
| Stunting |  |  |  |  |  |  |  |
| had |  | -.0256828 | .115381 | -0.22 | 0.824 | -.2518254 | .2004597 |
| agegroup |  | -.0276764 | .0614765 | -0.45 | 0.653 | -.1481681 | .0928152 |
| BWcateg |  | -.3826971 | .1153475 | -3.32 | 0.001 | -.608774 | -.1566202 |
| v190 |  | -.1327476 | .046673 | -2.84 | 0.004 | -.224225 | -.0412701 |
| v701 |  | -.0661625 | .0465119 | -1.42 | 0.155 | -.1573241 | .0249991 |
| HHsize |  | .0266355 | .1087494 | 0.24 | 0.807 | -.1865093 | .2397803 |

|  |  |  |  |  |  |  |  |  |
| --- | --- | --- | --- | --- | --- | --- | --- | --- |
| 293 | MBMI |  | .1126322 | .0854934 | 1.32 | 0.188 | -.0549318 | .2801962 |
| 294 | watersource |  | .008692 | .1552812 | 0.06 | 0.955 | -.2956536 | .3130376 |
| 295 | Tfacility |  | .0277633 | .1681797 | 0.17 | 0.869 | -.3018629 | .3573895 |
| 296 | Mothersage |  | -.3356838 | .1103067 | -3.04 | 0.002 | -.551881 | -.1194866 |
| 297 | Deliverly |  | .3040175 | .205598 | 1.48 | 0.139 | -.0989473 | .7069822 |
| 298 | Chbelow5 |  | -.8383208 | .383567 | -2.19 | 0.029 | -1.590098 | -.0865433 |
| 299 | DoBfeeding |  | .2369705 | .0660839 | 3.59 | 0.000 | .1074484 | .3664926 |
| 300 | boder |  | .1478164 | .0977222 | 1.51 | 0.130 | -.0437157 | .3393484 |
| 301 | m18n |  | -.0550137 | .1227062 | -0.45 | 0.654 | -.2955133 | .1854859 |
| 302 | v481 |  | .0007237 | .4333574 | 0.00 | 0.999 | -.8486412 | .8500887 |
| 303 | ANCtiming |  | .086621 | .0995567 | 0.87 | 0.384 | -.1085066 | .2817485 |
| 304 | NoANCvisists |  | -.1502899 | .1124919 | -1.34 | 0.182 | -.3707699 | .0701901 |
| 305 | v101 |  | .0280096 | .0132185 | 2.12 | 0.034 | .0021018 | .0539173 |
| 306 | v025 |  | -.1296497 | .146117 | -0.89 | 0.375 | -.4160337 | .1567343 |
| 307 | ReligioN |  | -.0494074 | .0511437 | -0.97 | 0.334 | -.1496472 | .0508324 |
| 308 | v106 |  | -.2041822 | .0900019 | -2.27 | 0.023 | -.3805828 | -.0277816 |
| 309 | v714 |  | -.052174 | .1393917 | -0.37 | 0.708 | -.3253768 | .2210288 |
| 310 | Depsode |  | .234093 | .1253 | 1.87 | 0.062 | -.0114905 | .4796764 |
| 311 | v467d |  | .2974141 | .1106 | 2.69 | 0.007 | .0806421 | .5141862 |
| 312 | _cons |  | .5521167 | .7207763 | 0.77 | 0.444 | -.8605789 | 1.964812 |

**Table 2:**
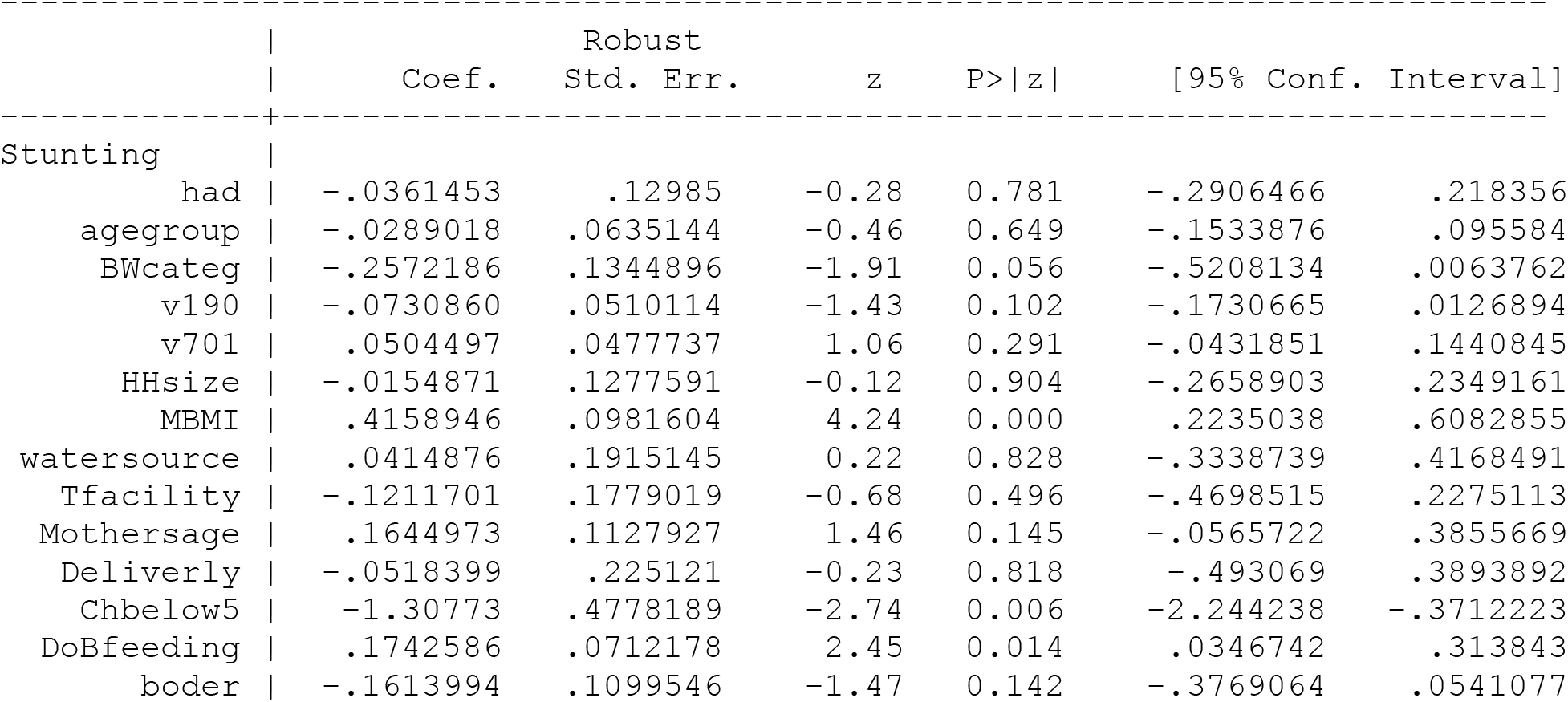
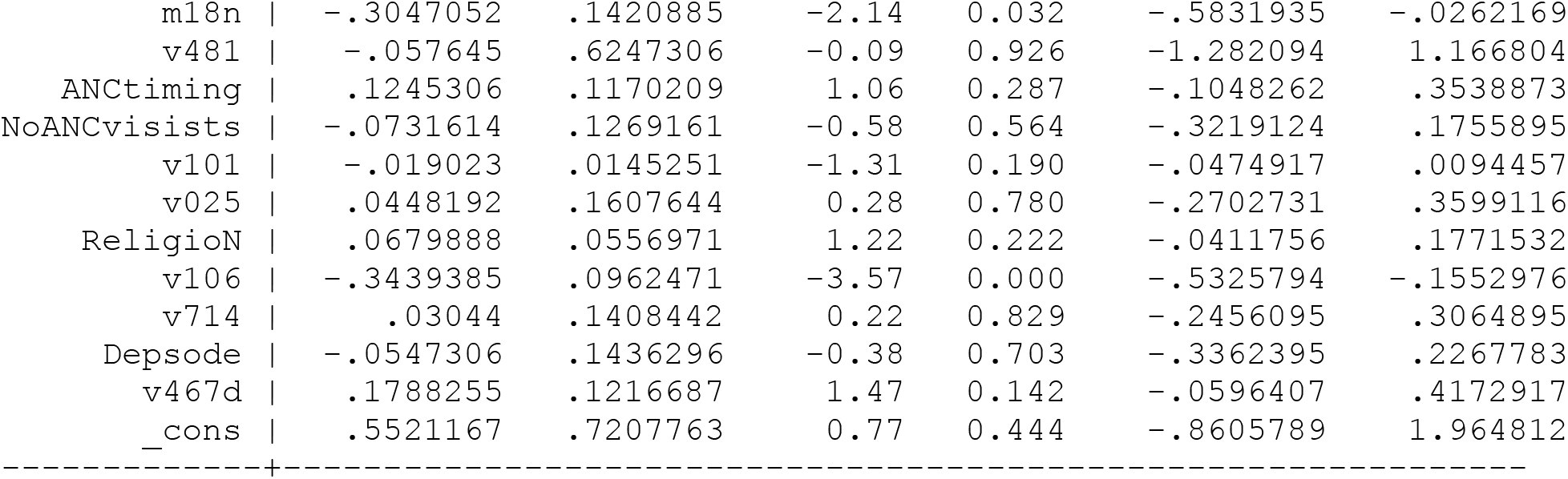
The risk factors responsible for stunting amongst the female under-fives (n=829)

|  |  |  | Robust |  |  |  |  |
| --- | --- | --- | --- | --- | --- | --- | --- |
|  |  | Coef. | Std. Err. | z | P>\|z\| | [95% Conf. Interval] |  |
| 332 | Stunting |  |  |  |  |  |  |
| 333 | had | -.0361453 | .12985 | -0.28 | 0.781 | -.2906466 | .218356 |
| 334 | agegroup | -.0289018 | .0635144 | -0.46 | 0.649 | -.1533876 | .095584 |
| 335 | BWcateg | -.2572186 | .1344896 | -1.91 | 0.056 | -.5208134 | .0063762 |
| 336 | v190 | -.0730860 | .0510114 | -1.43 | 0.102 | -.1730665 | .0126894 |
| 337 | v701 | .0504497 | .0477737 | 1.06 | 0.291 | -.0431851 | .1440845 |
| 338 | HHsize | -.0154871 | .1277591 | -0.12 | 0.904 | -.2658903 | .2349161 |
| 339 | MBMI | .4158946 | .0981604 | 4.24 | 0.000 | .2235038 | .6082855 |
| 340 | watersource | .0414876 | .1915145 | 0.22 | 0.828 | -.3338739 | .4168491 |
| 341 | Tfacility | -.1211701 | .1779019 | -0.68 | 0.496 | -.4698515 | .2275113 |
| 342 | Mothersage | .1644973 | .1127927 | 1.46 | 0.145 | -.0565722 | .3855669 |
| 343 | Deliverly | -.0518399 | .225121 | -0.23 | 0.818 | -.493069 | .3893892 |
| 344 | Chbelow5 | -1.30773 | .4778189 | -2.74 | 0.006 | -2.244238 | -.3712223 |
| 345 | DoBfeeding | .1742586 | .0712178 | 2.45 | 0.014 | .0346742 | .313843 |
| 346 | boder | -.1613994 | .1099546 | -1.47 | 0.142 | -.3769064 | .0541077 |

|  |  |  |  |  |  |  |  |  |
| --- | --- | --- | --- | --- | --- | --- | --- | --- |
| 347 | m18n |  | -.3047052 | .1420885 | -2.14 | 0.032 | -.5831935 | -.0262169 |
| 348 | v481 |  | -.057645 | .6247306 | -0.09 | 0.926 | -1.282094 | 1.166804 |
| 349 | ANCtiming |  | .1245306 | .1170209 | 1.06 | 0.287 | -.1048262 | .3538873 |
| 350 | NoANCvisists |  | -.0731614 | .1269161 | -0.58 | 0.564 | -.3219124 | .1755895 |
| 351 | v101 |  | -.019023 | .0145251 | -1.31 | 0.190 | -.0474917 | .0094457 |
| 352 | v025 |  | .0448192 | .1607644 | 0.28 | 0.780 | -.2702731 | .3599116 |
| 353 | ReligioN |  | .0679888 | .0556971 | 1.22 | 0.222 | -.0411756 | .1771532 |
| 354 | v106 |  | -.3439385 | .0962471 | -3.57 | 0.000 | -.5325794 | -.1552976 |
| 355 | v714 |  | .03044 | .1408442 | 0.22 | 0.829 | -.2456095 | .3064895 |
| 356 | Depsode |  | -.0547306 | .1436296 | -0.38 | 0.703 | -.3362395 | .2267783 |
| 357 | v467d |  | .1788255 | .1216687 | 1.47 | 0.142 | -.0596407 | .4172917 |
| 358 | _cons |  | .5521167 | .7207763 | 0.77 | 0.444 | -.8605789 | 1.964812 |
| 359 | -----+----- |  |  |  |  |  |  |  |

**Table 3:**
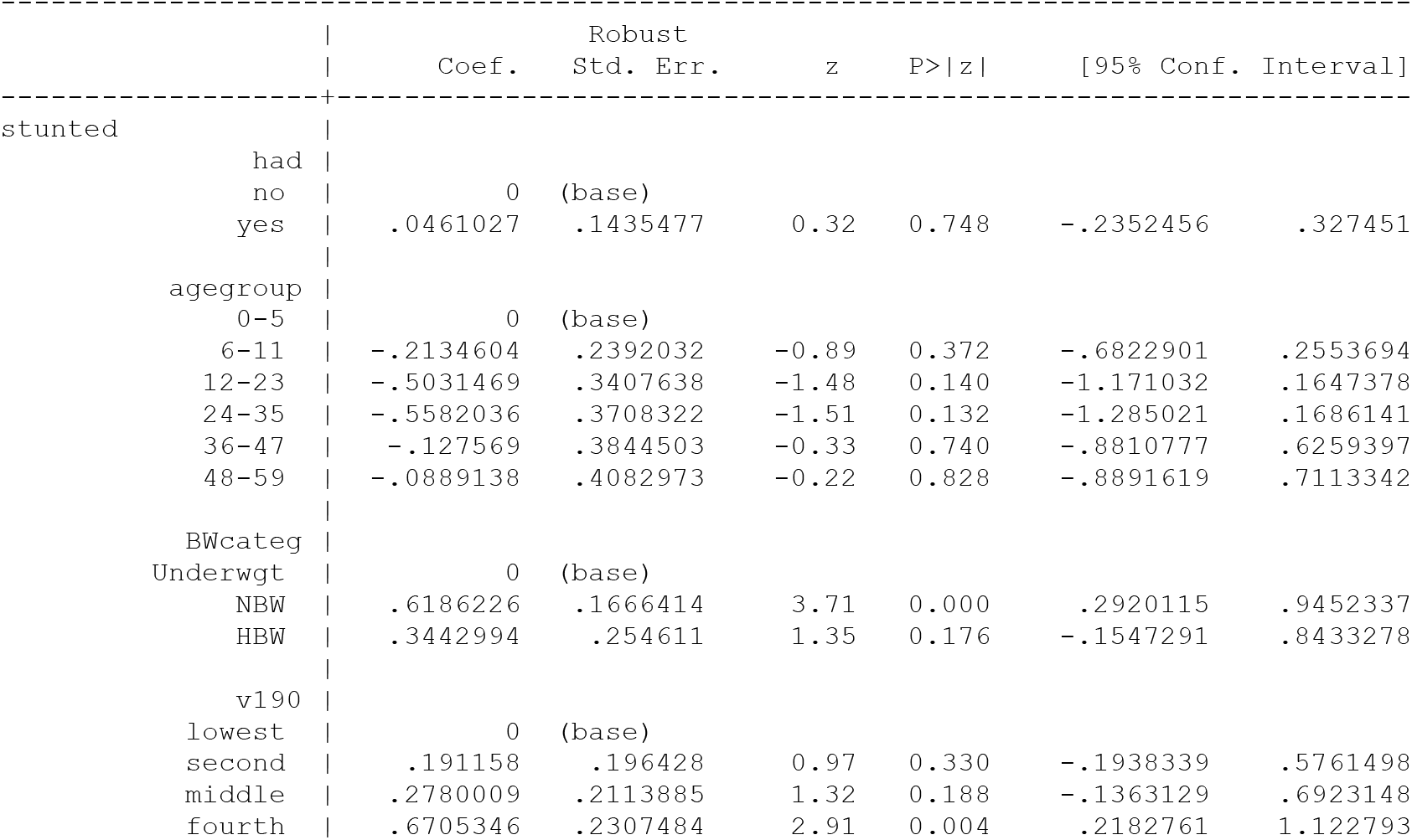
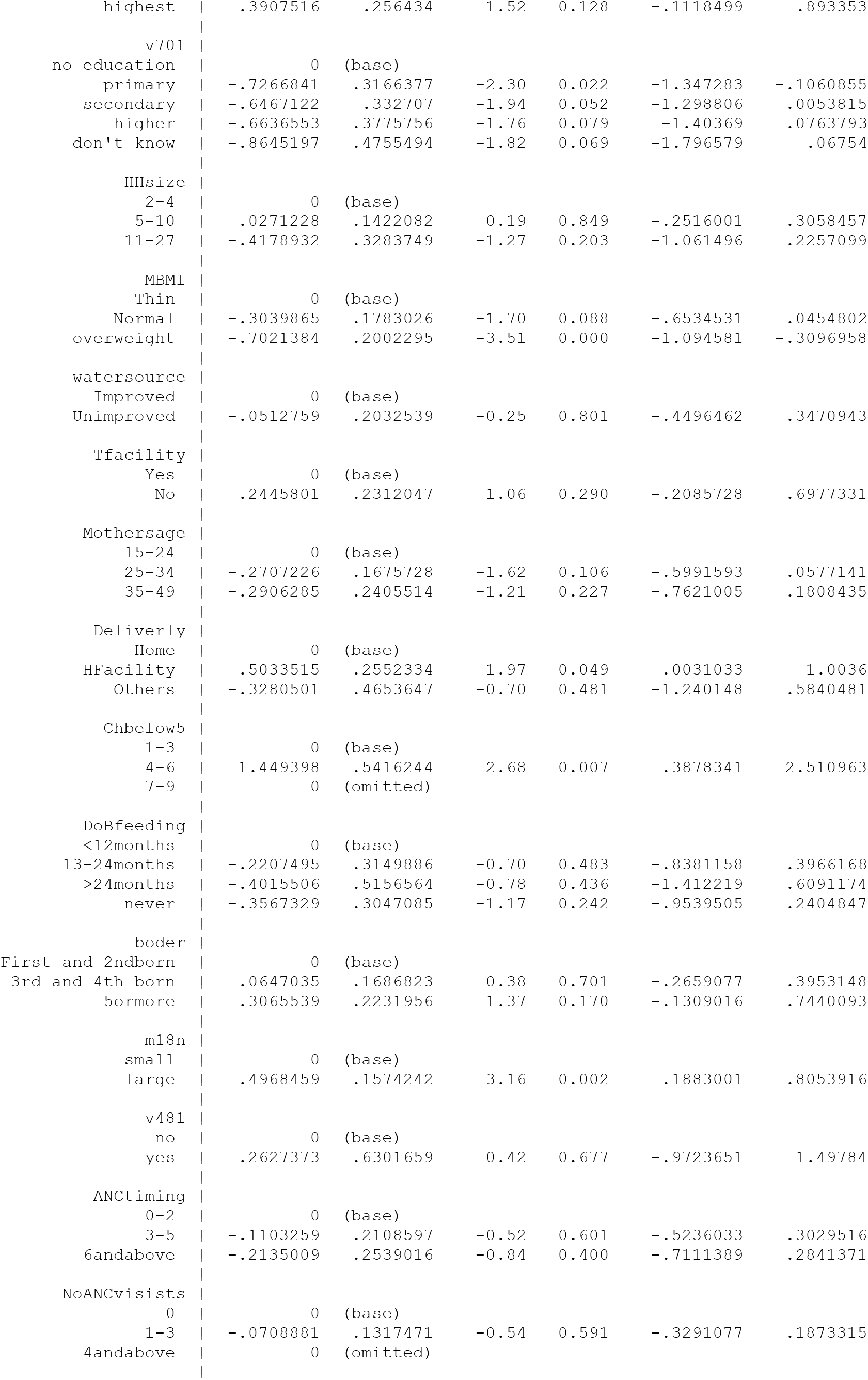
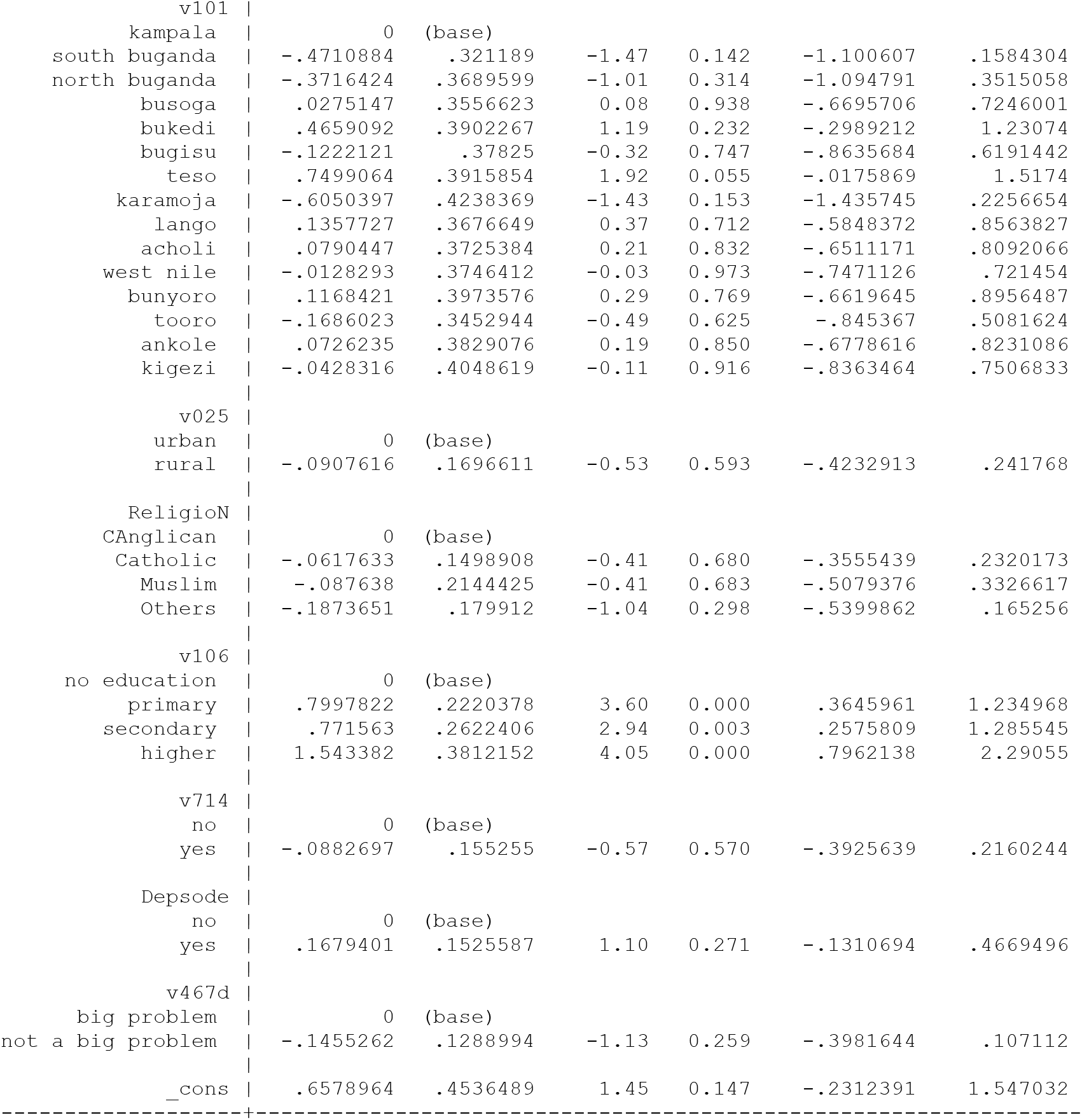
Determinants of stunting in male under-fives at factor level.

**Table 4:**
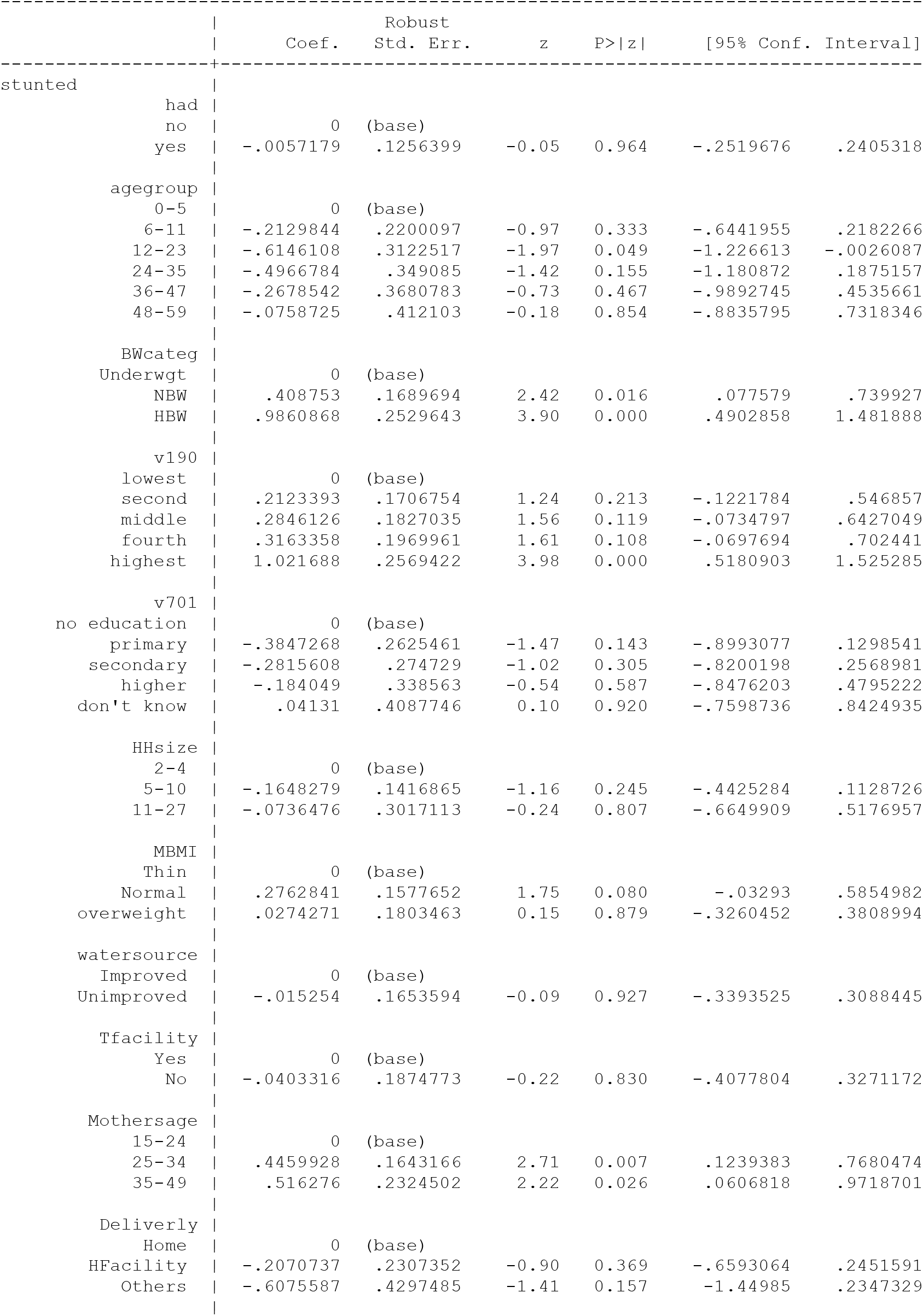
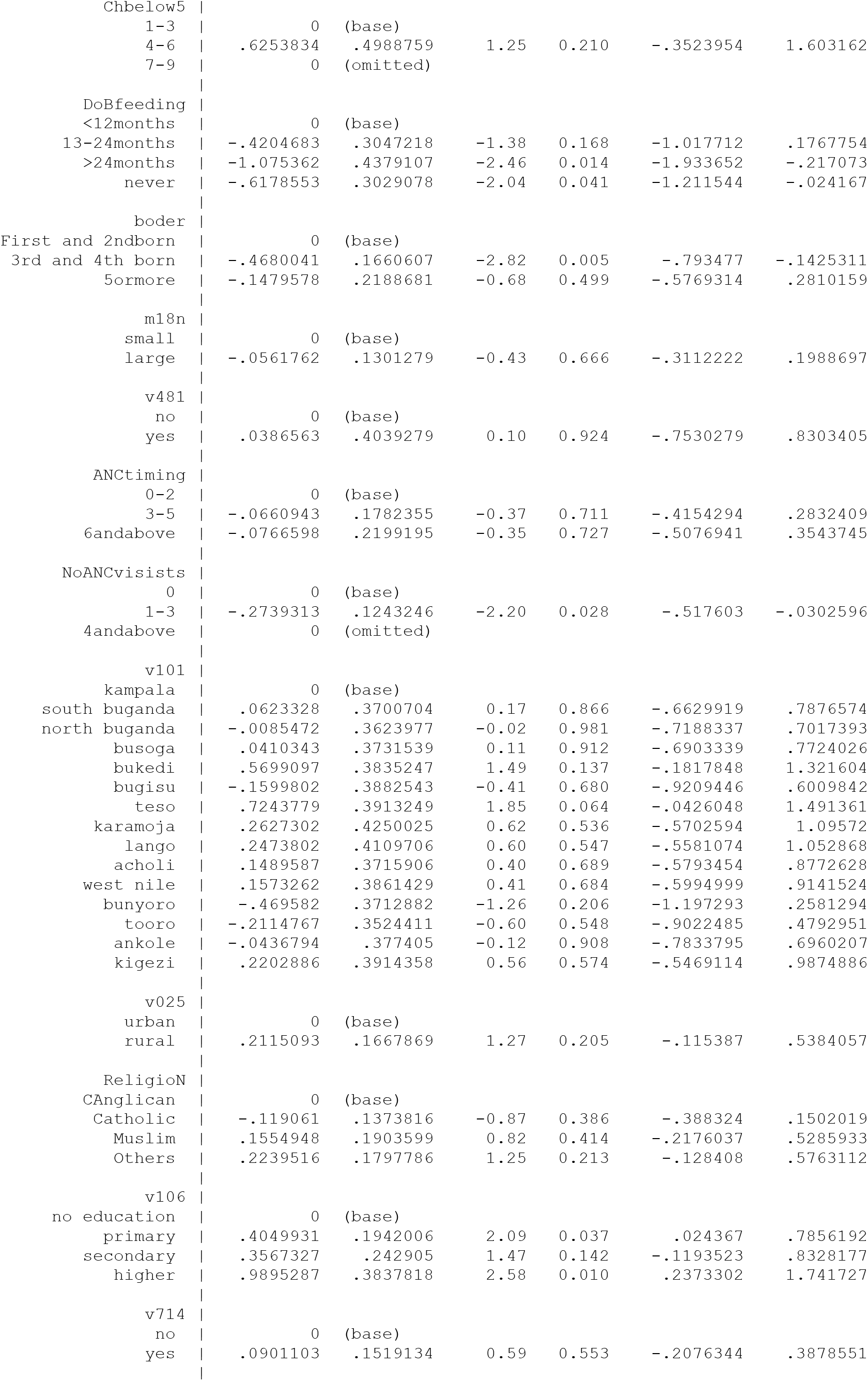
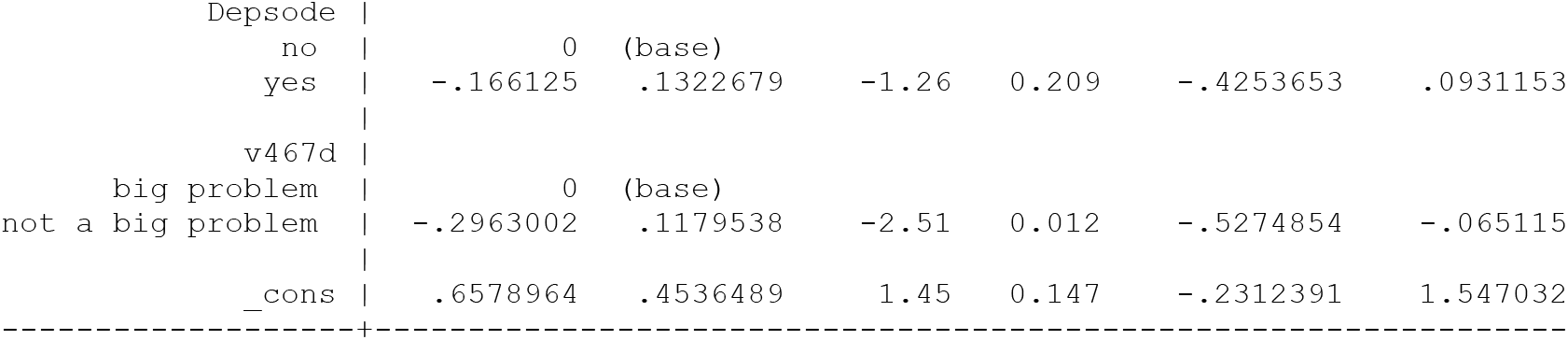
Determinants of stunting in female under-fives at factor level

Table 1 presents a model for the estimation of the risk factors that influence stunting with respect to the male child while Table 2 presents the risk factors that influence stunting with respect to the female child. It is observed that there is a variation among the risk factors that influence stunting amongst male and female children. The factors that were statistically significant in influencing stunting amongst the male children were not exactly the same factors influencing stunting amongst the female children. Overall, the most significant factors in both cases were; birthweight (BWcateg), number of children below 5years of age in the household, duration of breastfeeding and mother’s level of education. Household wealth index (v190), mother’s age, episodes of diarrhoea (Depsode), locality by region (v101) and distance to the nearest health center (v467d) were statistically significant in influencing stunting amongst the male under-fives but not amongst the female. Mother’s weight (MBMI) and the perceived size of children at birth were statistically significant in influencing stunting amongst the female under-fives but not amongst the male under-fives.

Table 1 and 2 also discloses that the household wealth index, weight of the child at birth, mother’s age, number of children below five years, education level of the mother and perceived size of the child at birth negatively influenced stunting while duration of breastfeeding, diarrhoea episodes and distance to the nearest health center positively influenced stunting of under-fives. Stunting being a serious health problem that affects both the physically and cognitively development of children which later affects their performance both at school and in the world of work needs to be tackled with respect to the gender or sex of the child. The strategies put in place to eliminated it should consider the differences in the risk factors that influence it with respect to the male and female differences in risk factors. It was necessary to make further analysis with respect to the factor levels of the variables in the model. Factor level analysis was crucial in identifying which categories within the factors were responsible for influencing stunting of under-fives in Uganda. As shown in Tables 3 and 4, the assumption that all the factor levels are equal in influencing stunting of under-fives in Uganda is put in the limelight. The first factor level for each factor was automatically selected as the base and consequently other factor levels were judged with reference to the selected or base factor level. The difference was considered statistically significant the base factor level if the p-value of a particular factor level was less than or equal to 0.1 that is, (p(z)<=10%))

At factor levels some of the variables which were not statistically significant proved to have some levels within themselves that tuned out to be statistically significant in influencing stunting. Age below 24 months of age for (agegroup) was significant from the base factor level for males and not for females in influencing stunting of under-fives. Birth weight and wealth index were still significant at all levels. Partners who were educated up to the level of primary were significant in influencing stunting for male while it was significant at all levels in influencing stunting for the female under-fives. Normal weight of mothers was significant in influencing stunting while for the female, it’s the overweight that was significant. For males, duration of breastfeeding was significant above 24 n=months while for female duration of breastfeed was not significant at all. Third and fourth birth rank were significant in influencing stunting in males it was not significant at all with respect to female under-fives. One to three antenatal visits were significant in influencing stunting with respect to male sex while for female sex, it was not significant at all levels. Mothers’ education level of primary and higher excluding secondary was significant in influencing stunting in male under-fives while it was significant at all factors with respect to influencing female stunting. Long distances to health center were a problem in influencing stunting among the male under-fives but was not the case with female under-fives. As far as regions are concerned, males are more a victim of stunting in Bukedi and Teso while female are more of a victim in Teso only. Being perceived to be large at birth was statistically significant in influencing stunting amongst the female under-fives but not amongst the male under-fives.

After the identification of significant variables that influence stunting in either male or female under-fives or both, it was prudent to identify the factors behind them in order to establish the mechanism of effect. Considered for the male under-fives were; weight of the child at birth, the household wealth index, presence of diarrhoea episodes, duration of breastfeeding and number of children below five years in the household. Theoretically, mother’s age group (mothers age), region and distance to the health center didn’t have influencing variables behind them. For the female group, weight at birth of the child, household wealth index, duration of breastfeeding, number of children below five years and mother’s and partners education level all having been found statistically significant influencers of stunting, were considered for path analysis. Figure 3 and 4 present the results for the male and the female groups respectively.

Figure 3 portrays the possible paths for statistically significant variables at various hierarchical levels with respect to the male under-fives. As it can be seen, at distal level, mother’s education level, household wealth index (v190), partner’s level of education (v701), mother’s working status and place of residence were outstanding influencers in the system. Tracking the roots of the paths in the system, made it possible to come up with various possible paths which would be key in dealing with the problem of stunting among the male under-fives in Uganda and elsewhere in the world. Among others, the most important possible paths identified were;

i. Through mother’s education level to influencing the number of under-fives in the households which influences the episodes of diarrhoea in an environment with poor toilet facilities and then stunting
ii. Low education level of the mother’s leads less employment opportunities which feeds into the household’s low wealth status which influences stunting either directly or indirectly through moderation of the duration of breastfeeding.
iii. Large household membership feeds into large number of children in the household that also influences the episodes of diarrhoea hence lowering the immunity system of the male under-fives leading to stunting.
iv. The education level of the partners feed into employments opportunities, then wealth status of the household and finally influences stunting either directly or through moderating the duration of breastfeeding.

Figure 4 was generated with respect to the female under-fives. Complete information was available for 829 cases for this path analysis. A close look and comparison of Figures 3 and 4 indicates differences in the possible paths as well as possible influencers of stunting amongst the under-fives (male and female). Among the distal factors that influence stunting for the female under-fives in the system were; mothers’ employment status, mother’s education level, residence, region, father’s education level and household wealth status. The most crucial paths identified in the system to be the possible channels through which stunting is influenced include;

i. The level of the mother through the mother’s weight, child’s weight and size at birth and finally stunting
ii. Through the mother’s education level to number of ante natal visits and timing to birthweight of the child and then stunting
iii. Education level of the mother influences stunting through wealth status of the household
iv. Employment status of the mother influences stunting of under-five females through its influence on the duration of breast-feeding.

## Discussions

Sex differences in growth paths and immune functions of the body, prenatally place boys at greater risk of infection and undernutrition, but these biological differences are interpreted by parents and within household contexts that are shaped by social and cultural norms which, in turn, influence care and feeding practices(29). Literature reviewed and cited in most sections of this study reveal that stunting affects the male and the female under-fives differently both in Uganda (17) and elsewhere in the world (30). It was thus worth to study and identify the risk factors and the causal mechanism that could explain the difference. The study was based on the modified 1991 UNICEF framework for child malnutrition which clearly categorizes and shows the interconnectivity of distal, intermediate, immediate and maternal level factors in a hierarchical order. The factors feed into each other implying a causal mechanism (Figure 1). This study made use of the structural equation modeling approach in its generalized form (GSEM) to identify not only the main risk factors responsible for stunting amongst the male and female groups but also the causal mechanism of the influencing risk factors with respect to the male and female groups.

Accordingly, this study clearly reveals that the risk factors which statistically influenced under-five stunting in Uganda differ with respect to the sex of the child. Male as a sex outcome was positively associated with stunting while being female was negatively associated with stunting of under-fives. Socioeconomic status of the household such as the household wealth status, education level of mother, mother’s employment status and residence turned out statistically significant influencers of in both male and female under-fives. Elsewhere for example in India, it was also observed that the risk factors that influenced stunting amongst male children differed from the risk factors that influenced stunting amongst the female children (31).

In this study, it was established that mother’s education level was a statistically significant risk factors in influencing stunting for both male and female children in Uganda which slightly differs from the findings from the rural primary school going children in India where mother’s education level predicted stunting amongst the female children only(31). The difference in the outcome may probably be because the school going children were somehow older than the children in our sample and also because our advanced modelling that generalizes the response to accommodate all types of variables. The current study also reveals the importance of education level in determining the wealth status of the household which is an indicator of wellbeing. This highlights the role of education in generating household wealth, information uptake and child care practices for maternal education and wealth greatly influence child development and stunting (32). Improving the wealth status of a household is key in reducing the problem of under-five stunting at household level but at a national level greater emphasis should be put on reducing the socioeconomic and particularly wealth and income inequality amongst the households (33). Overall, literature reveals that there is an inverse relationship between wealth status and level of stunting. The odds of being stunted reduces greatly with an increase in wealth index of the household (34). Lower wealth status of the household may also imply that there is little on table, the mothers nutrition intake may not be sufficient and as such they struggle with their children for the little food the mother eats (35).

Duration of breastfeeding was a significant factor in influencing stunting in both male and females amongst the under-fives in Uganda. This particular outcome echoes the recommendations of world health organization that emphasizes breastfeeding in the first six months of life () and the fact that infants who are exclusively breastfed for the first six months of their life have less morbidity and growth issues (36).

Age of the child was categorized into age groups of 0-5, 6-11, 12-23, 24-35, 36-47 and 48-59 months. The study reveals a statistically insignificant effect of agegroup in influencing stunting in both male and female children. Considering factor levels, the age group 12-23 was found statistically significant and different from other age groups for the female group. Prevalence of stunting increased with age whereby in most developing or low developed countries, the age-group 12–23-month was the most affected with more than 50% children being victims of stunting (37). Methodological differences may account for this contrast whereby most studies cited in literature used other methods that did not consider the generalized and hierarchical nature of variables. Other studies in Uganda (17) considered age categorizations that are too heterogenous in nature for example the age group “0-23” months and “24-59”. The attributes and nutrition requirements of children in these age-groups considered are obviously different. The study also shows that perceived size of the child at birth is statistically significant in influencing stunting amongst female under-fives but not amongst the male counterparts. Further scrutiny also reveals that there is no significant different among the factor levels with respect to the female under-five while it was revealed that male children whose size at birth was perceived large were significantly different from the rest of the groups (small and very large). Perceived size at birth was found to be statistically significant in influencing stunting among children whose perceived size was small and average size than their counterparts (38), however perceived size is relative and unclear and the results in this case are inconclusive.

The findings of this study reveal that mother’s age was a significant influencer of stunting amongst male under-fives but not amongst the female under-fives. The role of mother’s age in childcare cannot be taken lightly and this also is true with mother’s health (BMI) as well. This could be because young mothers may not have acquired enough technical know-how in taking care of children as well.

## Conclusion

The risk factors that influence stunting of under-fives with respect to the sex of the child was deeply analyzed in this study. Using GSEM modeling approach, the study paid attention to the family and link function for all the possible response factors in the model as well as establishing the causal mechanism of risk factors. Keen interest was put in identifying whether there were any differences in the set of factors that influence stunting amongst the male and female under-fives. It was clearly found out that indeed the set of factors that influence stunting amongst the male and female under-fives differ both in combination and in the chain or paths. The study recommends that the strategies to reduce stunting amongst the under-fives should factor in the sex or gender aspects of the under-fives. The findings of this study re-echo much of the key takes from the existing literature. Interventions to reduce stunting should consider the chain of influence and policy makers should be aware that the interventions that work for the male under-fives should be designed differently from the interventions to reduce stunting amongst the female under-fives.

## Data Availability

data for this publication can be accessed via the DHS programme: https://www.dhsprogram.com/publications/publication-fr333-dhs-final-reports.cfm

## Acknowledgement

The authors are so grateful to their respective institutions of higher learning for providing an enabling environment and time to have this work in place. We are grateful to African Center of Excellence in Data Science (ACE-DS), School of Economics, Gikondo campus at University of Rwanda for providing and enabling environment. We also extend our sincere thanks to the DHS programme for granting us permission and access to the UDHS data which has made this manuscript possible.

## Authors contribution

**Conceptualization:** Vallence Ngabo Maniragaba, Atuhaire Leonard and Rutayisire Peter Clever

**Methodology:** Vallence Ngabo, Atuhaire Leonard, Peter Clever Rutayisire.

**Formal analysis:** Vallence Ngabo Maniragaba

**Writing the draft:** Vallence Ngabo Maniragaba

**Review:** Atuhaire Leonard, Peter Clever Rutayisire

**Editing:** Vallence Ngabo Maniragaba

## Appendix 1 (over all GSEM model)

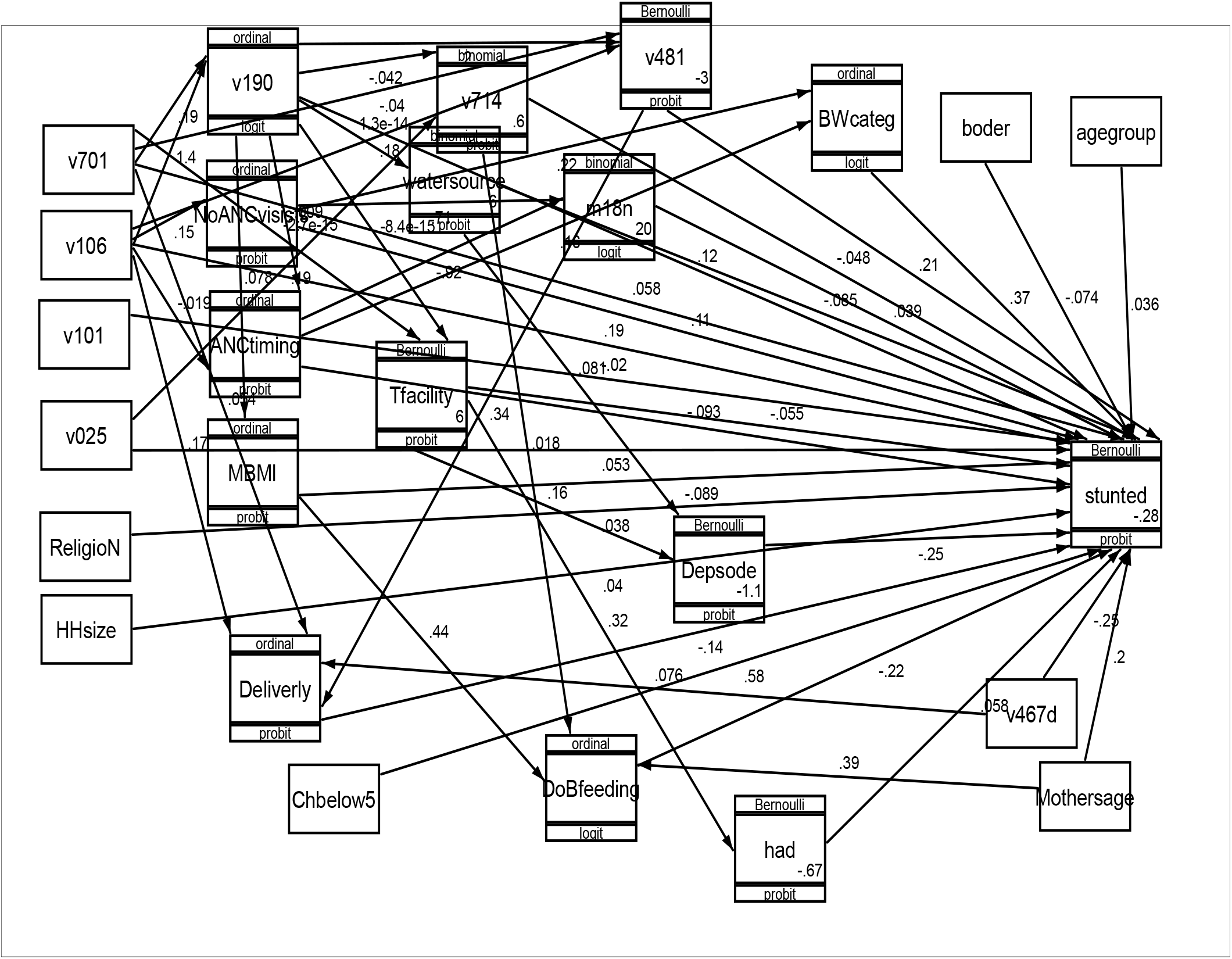

## References

1. De Onis M, Branca F. Childhood stunting: a global perspective. Matern Child Nutr. 2016;12:12–26.

2. Holden KR. Malnutrition and brain development: A review. Neurol Consequences Malnutrition. 2007;6:19.

3. Soekatri MYE, Sandjaja S, Syauqy A. Stunting was associated with reported morbidity, parental education and socioeconomic status in 0.5–12-year-old Indonesian children. Int J Environ Res Public Health. 2020;17(17):6204.

4. Prendergast AJ, Humphrey JH. The stunting syndrome in developing countries. Paediatr Int Child Health. 2014;34(4):250–65.

5. WHO. Title levels and trends in child malnutrition. 2020;

6. Wamani H, Åstrøm AN, Peterson S, Tumwine JK, Tylleskär T. Boys are more stunted than girls in sub-Saharan Africa: a meta-analysis of 16 demographic and health surveys. BMC Pediatr. 2007;7(1):1–10.

7. Teshome B, Kogi-Makau W, Getahun Z, Taye G. Magnitude and determinants of stunting in children underfive years of age in food surplus region of Ethiopia: the case of west gojam zone. Ethiop J Heal Dev. 2009;23(2).

8. Habaasa G. An investigation on factors associated with malnutrition among underfive children in Nakaseke and Nakasongola districts, Uganda. BMC Pediatr. 2015;15(1):1–7.

9. Bukusuba J, Kaaya AN, Atukwase A. Predictors of stunting in children aged 6 to 59 months: a case–control study in Southwest Uganda. Food Nutr Bull. 2017;38(4):542–53.

10. Olwedo MA, Mworozi E, Bachou H, Orach CG. Factors associated with malnutrition among children in internally displaced person\’s camps, northern Uganda. Afr Health Sci. 2008;8(4):244–52.

11. Ashaba S, Rukundo GZ, Beinempaka F, Ntaro M, LeBlanc JC. Maternal depression and malnutrition in children in southwest Uganda: a case control study. BMC Public Health. 2015;15(1):1–6.

12. Odei Obeng-Amoako GA, Karamagi CAS, Nangendo J, Okiring J, Kiirya Y, Aryeetey R, et al. Factors associated with concurrent wasting and stunting among children 6–59 months in Karamoja, Uganda. Matern Child Nutr. 2021;17(1):e13074.

13. Wamani H, Tylleskär T, Åstrøm AN, Tumwine JK, Peterson S. Mothers’ education but not fathers’ education, household assets or land ownership is the best predictor of child health inequalities in rural Uganda. Int J Equity Health. 2004;3(1):1–8.

14. Kasaye HK, Bobo FT, Yilma MT, Woldie M. Poor nutrition for under-five children from poor households in Ethiopia: Evidence from 2016 Demographic and Health Survey. PLoS One. 2019;14(12):e0225996.

15. Hagos S, Hailemariam D, WoldeHanna T, Lindtjørn B. Spatial heterogeneity and risk factors for stunting among children under age five in Ethiopia: A Bayesian geo-statistical model. PLoS One. 2017;12(2):e0170785.

16. Kimani-Murage EW, Muthuri SK, Oti SO, Mutua MK, Van De Vijver S, Kyobutungi C. Evidence of a double burden of malnutrition in urban poor settings in Nairobi, Kenya. PLoS One. 2015;10(6):e0129943.

17. Nankinga O, Kwagala B, Walakira EJ. Maternal employment and child nutritional status in Uganda. PLoS One. 2019;14(12):e0226720.

18. Boah M, Azupogo F, Amporfro DA, Abada LA. The epidemiology of undernutrition and its determinants in children under five years in Ghana. PLoS One. 2019;14(7):e0219665.

19. Vaivada T, Akseer N, Akseer S, Somaskandan A, Stefopulos M, Bhutta ZA. Stunting in childhood: an overview of global burden, trends, determinants, and drivers of decline. Am J Clin Nutr. 2020;112(Supplement_2):777S–791S.

20. Julianti E, Elni E. Determinants of stunting in children aged 12-59 months. Nurse Media J Nurs. 2020;10(1):36–45.

21. Rakotomanana H, Gates GE, Hildebrand D, Stoecker BJ. Determinants of stunting in children under 5 years in Madagascar. Matern Child Nutr. 2017;13(4):e12409.

22. Chirande L, Charwe D, Mbwana H, Victor R, Kimboka S, Issaka AI, et al. Determinants of stunting and severe stunting among under-fives in Tanzania: evidence from the 2010 cross-sectional household survey. BMC Pediatr. 2015;15(1):1–13.

23. Tiwari R, Ausman LM, Agho KE. Determinants of stunting and severe stunting among under-fives: evidence from the 2011 Nepal Demographic and Health Survey. BMC Pediatr. 2014;14(1):1–15.

24. Uganda Bureau of Statistics (UBOS) and ICF. Uganda Demographic and Health Survey 2016. Kampala, Uganda and Rockville, Maryland, USA; 2018.

25. Group Whomgrs, de Onis M. WHO Child Growth Standards based on length/height, weight and age. Acta Paediatr. 2006;95:76–85.

26. Yerushalmy J. The classification of newborn infants by birth weight and gestational age. J Pediatr. 1967;71(2):164–72.

27. Lombardi S, Santini G, Marchetti GM, Focardi S. Generalized structural equations improve sexual-selection analyses. PLoS One. 2017;12(8):1–20.

28. Workie DL, Tesfaw LM. Bivariate binary analysis on composite index of anthropometric failure of under-five children and household wealth-index. BMC Pediatr. 2021;21(1):1– 13.

29. Thompson AL. Greater male vulnerability to stunting? Evaluating sex differences in growth, pathways and biocultural mechanisms. Ann Hum Biol. 2021;48(6):466–73.

30. Thurstans S, Opondo C, Seal A, Wells J, Khara T, Dolan C, et al. Boys are more likely to be undernourished than girls: a systematic review and meta-analysis of sex differences in undernutrition. BMJ Glob Heal. 2020;5(12):e004030.

31. Pal S, Bose K. Prevalence and Sex Specific Determinants of Stunting among Rural Primary School Children of Hooghly District, West Bengal, India. Int J Stat Sci. 2020;19:67–86.

32. Efevbera Y, Bhabha J, Farmer PE, Fink G. Girl child marriage as a risk factor for early childhood development and stunting. Soc Sci Med. 2017;185:91–101.

33. Rizal MF, van Doorslaer E. Explaining the fall of socioeconomic inequality in childhood stunting in Indonesia. SSM-population Heal. 2019;9:100469.

34. Pillai VK, Maleku A. Women’s education and child stunting reduction in India. J Soc Soc Welf. 2019;46:111.

35. Hien NN, Hoa NN. Under three years of age in nghean, Vietnam. Pakistan J Nutr. 2009;8(7):958–64.

36. Organization WH. The optimal duration of exclusive breastfeeding: a systematic review. 2001;

37. Marriott BP, White A, Hadden L, Davies JC, Wallingford JC. World Health Organization (WHO) infant and young child feeding indicators: associations with growth measures in 14 low-income countries. Matern Child Nutr. 2012;8(3):354–70.

38. Akombi BJ, Agho KE, Hall JJ, Merom D, Astell-Burt T, Renzaho A. Stunting and severe stunting among children under-5 years in Nigeria: A multilevel analysis. BMC Pediatr. 2017;17(1):1–16.

